# Incubation-period estimates of Omicron (BA.1) variant from Taiwan, December 2021–January 2022, and its comparison to other SARS-CoV-2 variants: a statistical modeling, systematic search and meta-analysis

**DOI:** 10.1101/2023.07.20.23292983

**Authors:** Andrei R. Akhmetzhanov, Hao-Yuan Cheng, Jonathan Dushoff

**Author notes:** Corresponding author: Andrei R. Akhmetzhanov, PhD Institute of Epidemiology and Preventive Medicine, College of Public Health, National Taiwan University; Rm. 541, No.17 Xuzhou Rd. Zhongzheng District, Taipei 10055, Taiwan.

## Abstract

**Background:** The ongoing COVID-19 pandemic has seen several variants of concern, including the Omicron (BA.1) variant which emerged in October 2021. Accurately estimating the incubation period of these variants is crucial for predicting disease spread and formulating effective public health strategies. However, existing estimates often conflict because of biases arising from the dynamic nature of epidemic growth and selective inclusion of cases. This study aims to accurately estimate of the Omicron (BA.1) variant incubation period based on data from Taiwan, where disease incidence remained low and contact tracing was comprehensive during the first months of the Omicron outbreak.

**Methods:** We reviewed 100 contact-tracing records for cases of the Omicron BA.1 variant reported between December 2021 and January 2022, and found enough information to analyze 70 of these. The incubation period distribution was estimated by fitting data on exposure and symptom onset within a Bayesian mixture model using gamma, Weibull, and lognormal distributions as candidates. Additionally, a systematic literature search was conducted to accumulate data for estimates of the incubation period for Omicron (BA.1/2, BA.4/5) subvariants, which was then used for meta-analysis and comparison.

**Results:** The mean incubation period was estimated at 3.5 days (95% credible interval: 3.1–4.0 days), with no clear differences when stratified by vaccination status or age. This estimate aligns closely with the pooled mean of 3.4 days (3.0–3.8 days) obtained from a meta-analysis of other published studies on Omicron subvariants.

**Conclusions:** The relatively shorter incubation period of the Omicron variant, as compared to previous SARS-CoV2 variants, implies its potential for rapid spread but also opens the possibility for individuals to voluntarily adopt shorter, more resource-efficient quarantine periods. Continual updates to incubation period estimates, utilizing data from comprehensive contact tracing, are crucial for effectively guiding these voluntary actions and adjusting high socio-economic cost interventions.

## 1. Introduction

The emergence of the Omicron variant in South Africa in October 2021 marked a remarkable shift in the trajectory of COVID-19 pandemic [1,2]. Compared to preceding variants, Omicron displayed higher transmissibility but lower severity [3–6]. This, coupled with adequate vaccination coverage and pandemic fatigue, led to the relaxation of most non-pharmaceutical interventions and the discontinuation of zero-COVID policy, culminating in the declaration of the pandemic end on 5 May 2023 [7,8]. With this altered COVID-19 epidemiology and the downgrading of COVID-19 status [9,10], limiting epidemiological investigations, the accurate assessment of Omicron variant parameters has become increasingly challenging.

Contrasting many countries, Taiwan effectively contained the early spread of the Omicron (BA.1) variant. Active case finding, contact tracing, quarantine of all close contacts, and isolation of confirmed cases maintained daily COVID-19 counts to remain below 20-30 from December 2021 to April 2022. Particularly in the initial two months of the outbreak, successful investigation of cases’ infection source and mandatory RT-PCR testing for all identified close contacts likely resulted in high case ascertainment. This provided a unique opportunity to gather exposure data and precisely estimate the incubation period distribution, a critical epidemiological parameter. However, a much larger wave of Omicron infections, associated with the predominant BA.2 subvariant, emerged in Taiwan in April 2022 [11,12].

One of the earliest published estimates proposed a mean Omicron (BA.1) incubation period of 3.2 days (95% confidence interval: 2.9-3.6 days; based on 258 cases) [13]. Later studies indicated slightly longer, but relatively consistent estimations: 3.5 days (3.2-3.8 days; 80 cases) for a study in Italy [14], 3.6 days (3.5-3.6 days; 2682 cases) from a large-cohort study in France [15], and 3.8 days (3.5-4.1 days; 114 cases) for a study in China [16]. The systematic review and meta-analysis by Wu et al. suggested an overall pooled mean of 3.4 days (2.9-4.0 days) [17]. However, these estimates were contested by Park et al., who argued that bias could arise from the dynamic nature of epidemic growth if the data are collected during its escalating phase [18]. Such circumstances might overrepresent cases with recent infection history, thereby favoring shortened incubation periods. Upon revising the initial estimate [13], they proposed a mean of 4.2 days (3.6-4.9 days).

Conversely, recent studies by Tanaka and colleagues reported comparatively shorter mean estimates of 3.1 days (2.1-4.1 days; 77 cases) [19] and 2.9 days (2.6-3.2 days; 68 cases) [20]. These authors, as does the present study, relied on contact tracing data collected in Ibaraki prefecture, Japan. However, their results may have been influenced by a selection bias as they included only cases with a one-day exposure period, which favored shorter incubation periods, as discussed in [21]. Intriguingly, two other studies [22,23] also applied the same case inclusion criteria and estimated a mean closely mirroring the results of Tanaka and colleagues, but notably shorter than others. The recently published study from Singapore [22] estimated the mean at 2.8 days (0.8–7.0 days; 36 cases) and an independent internal report of the National Institute of Infectious Diseases, Japan [23] stated the mean at 2.9 days (2.6-3.2 days; 35 cases).

In contrast, our dataset from Taiwan is likely less influenced by the aforementioned biases. Given the steady, low incidence of COVID-19 in Taiwan between December 2021 and January 2022, the potential impact of the dynamic bias highlighted by Park et al. is likely negligible. Furthermore, our analysis incorporates cases with exposure windows wider than a single day, reducing potential selection bias. Our dataset provides a unique opportunity to estimate the incubation period under altered conditions, offering a basis for comparison with previous studies and deriving potentially unbiased estimates of the incubation period.

As the COVID-19 pandemic progresses, the emergence of new, more transmissible variants such as Omicron BA.2, BA.4/5, XBB.*, emerged later in 2022-2023, complicates the task of accurately estimating epidemiological parameters, including the incubation period. This challenge is more profound in regions with widespread disease, where various potential sources of infection complicate the identification of specific exposure times. This study estimates the incubation period of Omicron (BA.1) variant by effectively leveraging highly accurate contact tracing data, thus minimizing the influence of several potential biases. The precise estimation of the incubation period continues to be a critical challenge in epidemiological research, as the accurate assessment of COVID-19 transmission remains essential for informing public health policy.

## 2. Methods

### 2.1. Data collection

Data used in this study were obtained through the analysis of daily public and internal epidemiological reports from the Taiwan Centers for Disease Control (CDC) and press conferences of the Central Epidemic Command Center (CECC) [24,25]. All records were obtained from epidemiological investigations conducted by local authorities in Taiwan and the Taiwan CDC.

The study considered all cases reported from 27 December 2021, the date of the first report of the local Omicron case in Taiwan, until 18 January 2022 were considered for inclusion in the analysis. All data records were initially compiled by H.-Y.C. and subsequently cross-verified by A.R.A.

Out of 128 cases reported during this period, one case confirmed with Delta variant and 27 cases with missing data on the date of symptom onset were excluded from our study. Among remaining 100 cases, 94 were genetically sequenced and identified with the Omicron BA.1.1.529 variant, and 6 could be epidemiologically linked to a case with a known infection of the Omicron variant.

The data, being interval-based, set constraints on exposure, *e*_*i*_, and symptom onset time, *o*_*i*_, such that *E*_*L*,*i*_ ≤ *e*_*i*_ ≤ *E*_*R*,*i*_ and *O*_*L*,*i*_ ≤ *o*_*i*_ ≤ *O*_*R*,*i*_ for each case *i*. Before including a case in the study, we made the following adjustments: (1) symptom onset dates were all identified by a one-day length (*O*_*R*,*i*_ ≔ *O*_*L*,*i*_ + 1 *day*); (2) if the right boundary of the exposure interval, *E*_*R*,*i*_, was unknown or established later than the right boundary of the symptom onset interval, *O*_*R*,*i*_, it was assigned to that boundary (i.e., *E*_*R*,*i*_ ≔ *O*_*R*,*i*_ if *E*_*R*,*i*_ is unknown or *E*_*R*,*i*_ > *O*_*R*,*i*_); (3) if the case lacked other exposure information except that its right boundary aligns with the right boundary of symptom onset time interval (*E*_*R*,*i*_ ≡ *O*_*R*,*i*_),and the left boundary was unknown, it was excluded from the analysis for being non-informative for the estimation. Out of 70 cases finally included in the study, 65 had definitive exposure time intervals and 5 cases were left-censored (**Figure 1**).

**Figure 1:**
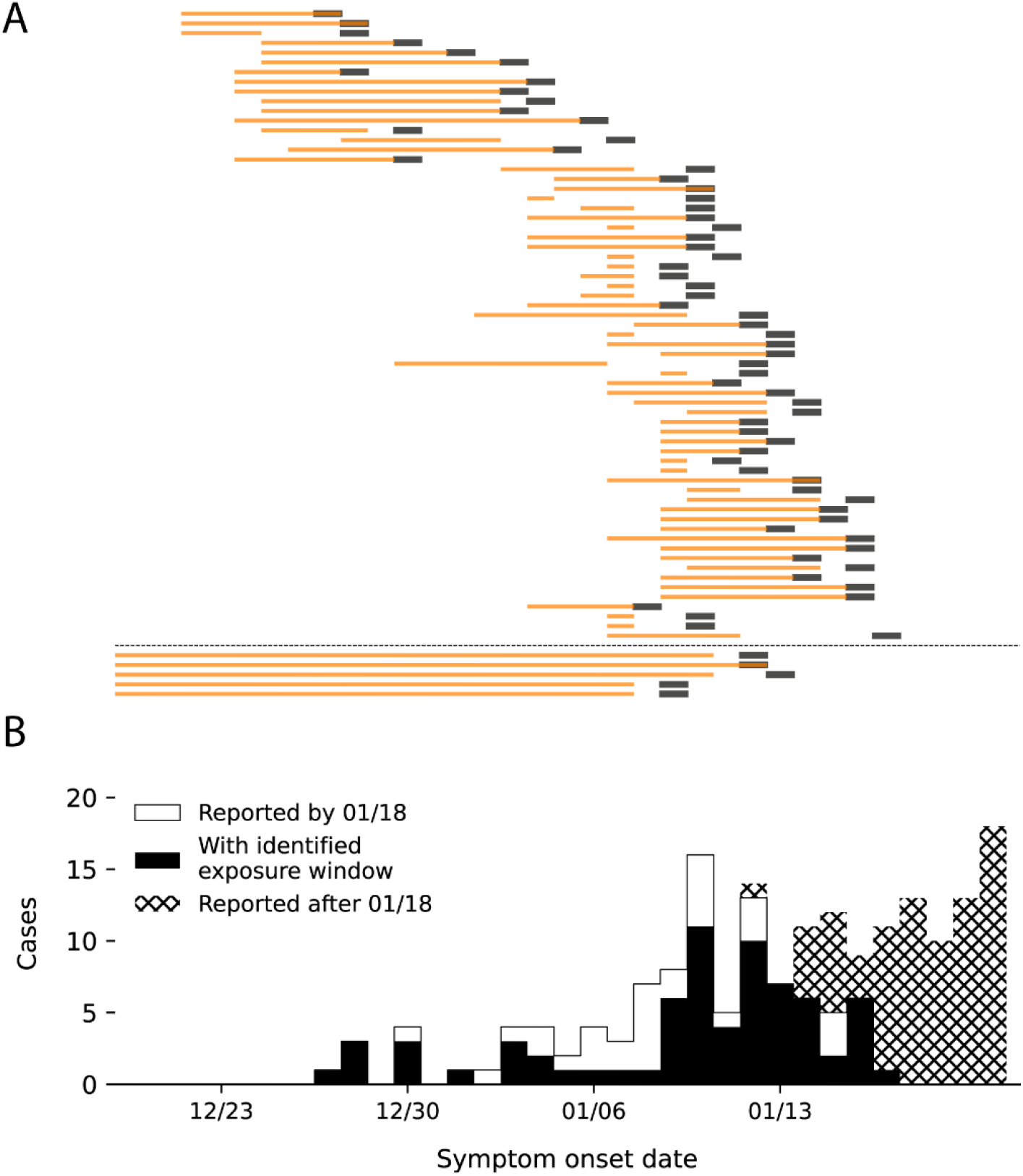
Data on confirmed local COVID-19 cases associated with Omicron BA.1 variant that was collected in Taiwan from 25 December 2021 through 18 January 2022. **(A)** shows the exposure windows (in orange) and symptom onset days (in black) for 70 cases included in our study. The dashed line separates 65 cases with a definitive exposure window (above) and 5 cases with a left-censored exposure window (below). **(B)** shows the epidemiological curve for symptomatic COVID-19 cases by their date of symptom onset. White bars indicate all cases considered for inclusion, black bars indicate the included cases, while the hatched bars indicate excluded cases as they were reported after the cut-off date of January 18, 2022. The horizontal axis is shared among both panels (A) and (B).

### 2.2. Statistical framework

To ascertain the distribution of the incubation period, we employed a Bayesian mixture model, fitting the data using three distributions: gamma, Weibull, or lognormal. Each distribution was given a relative weight within the model. The mean, *m*_*inc*_, and standard deviation (SD), *s*_*inc*_, of the incubation period were kept common across all three distributions to promote better convergence of the Markov chain Monte Carlo (MCMC) simulations [26].

The likelihood was given by a sum of the three component likelihoods, each weighted by *w*_*l*_:

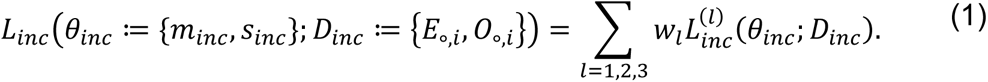

Here ∘≔ {*L*, *R*} and ∑_*l*_ *w*_*l*_ = 1. The component likelihoods were doubly censored [27,28] and right truncated at *T*_*inc*_ = 18 *January* 2022:

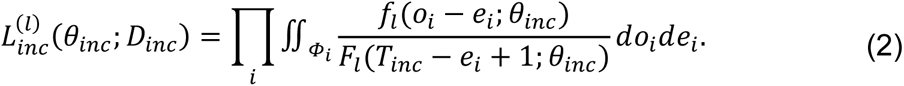

We assumed *e*_*i*_ and *o*_*i*_ to be randomly uniformly distributed within their intervals, defining the respective area in the state space as:

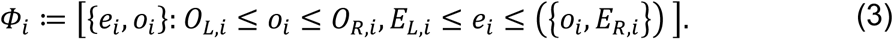

The function *f*_*l*_(.; *θ*_*inc*_) represents the probability density function for distribution *l* (*l* = 1,2,3), while the function *F*_*l*_(.; *θ*_*inc*_) is the cumulative distribution function of *f*_*l*_.

Finally, the posterior probability for selecting the distribution *l* was defined by the expression:

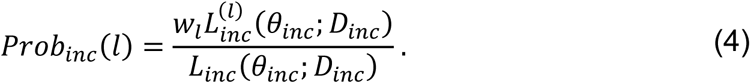

The estimated parametric distribution of the incubation period was then validated by comparing with the empirical cumulative distribution function (ECDF). The Bayesian model allowed to infer the posteriors for individual exposure and symptom onset times of the cases, {*e*_*i*_, *o*_*i*_}. Then the ECDF, *F*_*emp*_(*t*), was estimated by a fraction of records with time intervals (*o*_*i*_ − *e*_*i*_) shorter than a given time moment *t*.

### 2.3. Systematic search and meta-analysis

To collect data on estimation of the incubation period for Omicron subvariants, we conducted a literature search on 14 July 2023 using the PubMed database with the following search inquiry: “(Omicron) AND ((Incubation) OR (generation time) OR (serial interval)) AND LitCTRANSMISSION[filter]”. The query was designed to identify Omicron-related publications from a collection consistent with COVID-19 transmission literature gathered by PubMed (“LitCTRANSMISSION”). To ensure a broad range of studies was included, we incorporated two additional epidemiological parameters, the generation time and serial interval, in our search as some studies aimed to estimate these estimates but also reported the estimates of the incubation period [14].

Out of 110 references obtained from the search, 85 were deemed irrelevant upon manual inspection (**Figure 2**). 13 references were excluded from our meta-analysis for various reasons: eight were written not in English, precluding a reliable assessment of their methodologies; two investigated the incubation period but did not explicitly state the estimates; one study [29] was not peer-reviewed at the time of search and it was used only in our discussion; two references [17,30] were previously published systematic search and meta-analysis studies, their pooled means were used for comparative purposes.

**Figure 2:**
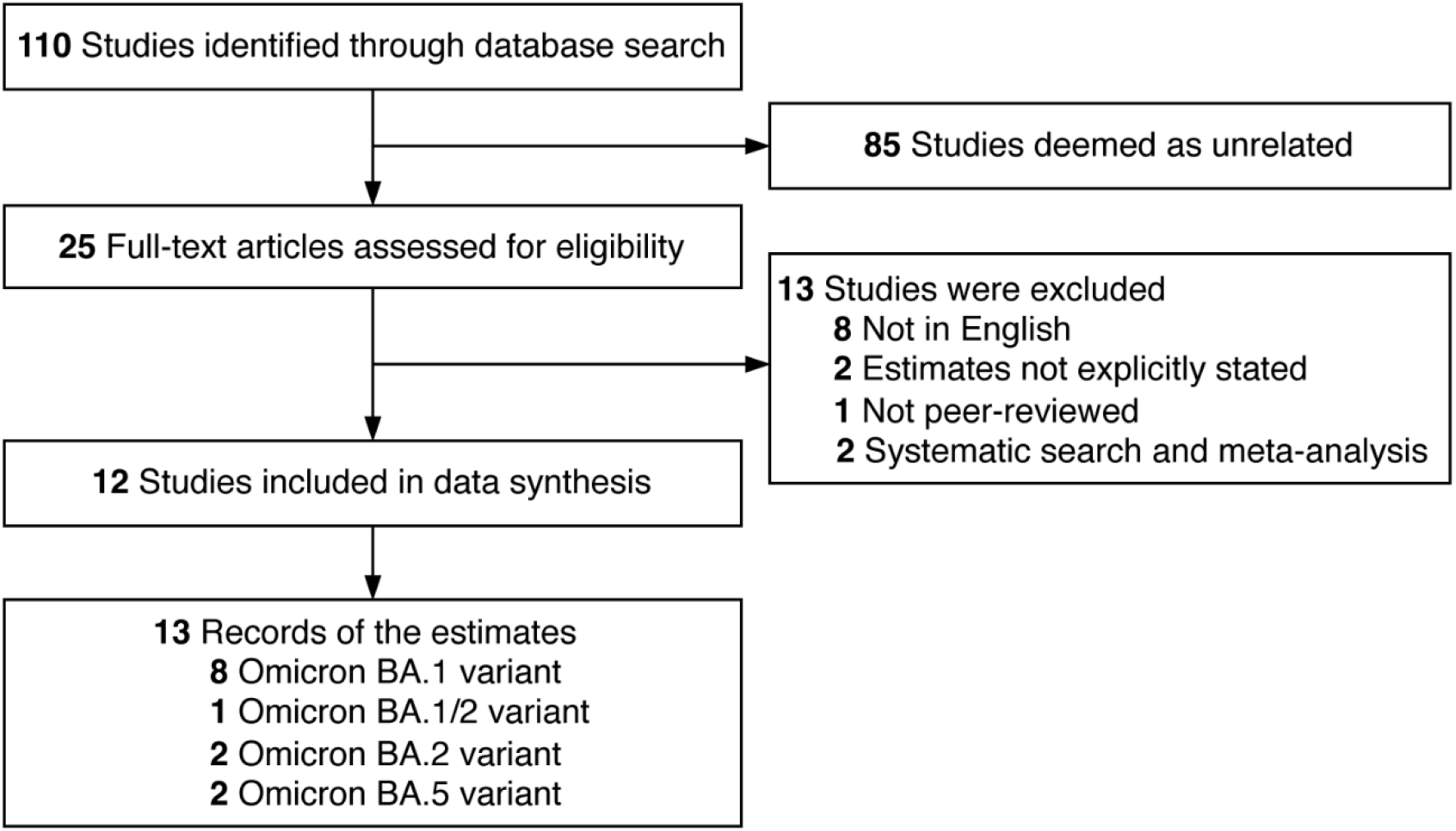
Flow diagram described how we identified previously published studies on estimates of incubation period for Omicron variants.

The remaining 12 references provided estimates of the incubation period for Omicron (BA.1, BA.2, BA.5) variants. Eight of these references [13,14,15,16,18,19,20,22] reporting the estimates of the incubation period for Omicron BA.1 variant were used to derive the pooled mean. One reference [31] provided an estimate for a mix of BA.1 and BA.2 cases, two references [32,33] provided an estimate for BA.2, and two references [20,34] reported an estimate for BA.5. These were used for visual comparison with our BA.1 estimates and as an update of earlier reviews [17,30].

### 2.4. Technical details

Incubation period distribution and meta-analysis were performed using Markov Chain Monte Carlo (MCMC) sampling techniques. Bayesian estimation was implemented using Stan software [35]. Each run of simulations was consistent of 4 parallel chains with 15,000 posterior draws including 2,500 draws used for tuning-in and disregarded for the final output.

## 3. Results

We analyzed **70 case records** to determine the incubation period with an average patient age of 35.3 years (range 1–66 years), and with females constituting 62% (43 cases) of the cohort. The vaccination status of 13 cases (18.6%) was unknown, while 41 case (58.6%) were breakthrough infections, with majority having received two doses of the ChAdOx1 nCoV-19 vaccine (AZD1222) developed by AstraZeneca, U.K. [35]. The mean age of vaccinated individuals was 40.1 years (range: 17–66) and of non-vaccinated individuals was 22.6 years (range: 1–57). The younger age distribution for non-vaccinated group reflects their partial ineligibility for vaccination, as only over 12 years of age were eligible for vaccination in Taiwan by mid-2022.

The estimated distribution of the incubation period and its comparison with the pre-Alpha (ancestral) variant [28] is shown in **Figure 3A**. The estimated mean was at 3.51 days (95% credible interval [CrI]: 3.06–3.99 days) and the standard deviation (SD) at 1.23 days (95% CrI: 0.85–1.82), with the 95th percentile mean posterior at 5.75 days. The lognormal distribution was most likely to be selected among the investigated distributions, with posterior mean of 68%. The parametric cumulative distribution function (CDF) closely mirrored the empirical CDF (**Supplementary Figure 1A**). Upon stratifying the incubation period by vaccination status or age, no significant differences were noted (**Supplementary Figure 1BC**), although non-vaccinated or older adults (>50 years of age) tended to have a longer incubation period. Our meta-analysis revealed a pooled mean of 3.40 days (95% CrI: 2.96–3.84 days), closely aligning with our estimate (**Figure 3B**). This pooled mean was also consistent with two other previously published meta-analyses [17,30] (**Figure 3C**).

**Figure 3:**
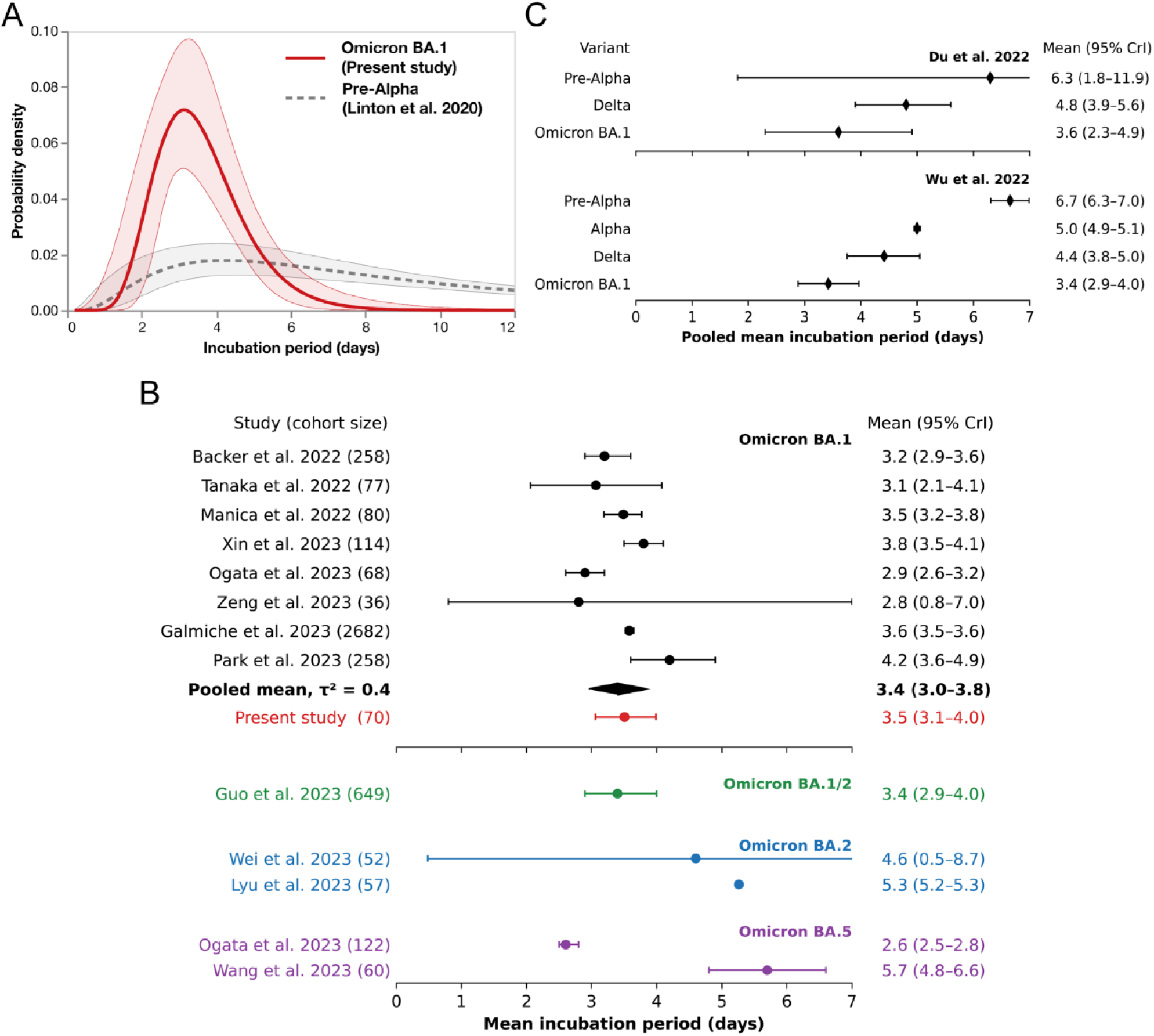
**(A)** shows the estimated incubation period distribution for the Omicron BA.1 variant compared to the earlier estimate for the pre-Alpha (ancestral) variant [28]. **(B)** shows the forest plot for the meta-analysis of mean incubation periods for Omicron (BA.1, BA.2, BA.5) subvariants. The dot indicates the posterior mean, while whiskers indicate 95% credible interval (CrI). The pooled mean is indicated in bold, while the estimate of the present study is indicated in red and it was not a part of the meta-analysis. **(C)** shows the comparison of different pooled means across various SARS-CoV-2 variants selected from two meta-analyses (Du et al. [30] and Wu et al. [17]).

## 4. Discussion

In our study, we estimated the incubation period distribution for Omicron BA.1 infections, yielding a mean of 3.51 days (95% confidence interval: 3.06–3.99 days, SD: 1.23 days). The consistency of our mean with the pooled means from relevant, up-to-date publications (as of 14 July 2023) and two prior meta-analyses underscore its credibility. These results stem from collected in Taiwan between 25 December 2021 and 18 January 2022, a period during which the daily occurrence of COVID-19 was less than 20 cases and the epidemic was not in an escalating phase. The valuable data resulted from the rigorous active case finding and contact tracing efforts of Taiwan’s local agencies and CDC. Given that the contact tracing teams were not overwhelmed during that period, it is plausible that the data were highly ascertained.

Compared to the estimated incubation period of previous variants of SARS-CoV2, the relatively shorter incubation period of Omicron variant may suggest a faster spread; On the other hand, a short incubation period also suggests a short quarantine period may already enough for containing the outbreak and save more resources. Keeping updating the estimates of incubation period using the updated data acquired from comprehensive contact tracing helps us more efficiently adjust the interventions with high socio-economical cost.

Besides, our analysis did not reveal a substantial difference in incubation periods between vaccinated and non-vaccinated individuals. Notably, the majority of vaccinated cases had received the ChAdOx1 nCoV-19 vaccine (AZD1222), which has been reported to demonstrate limited protection against symptomatic disease caused by Omicron variant [36]. Furthermore, we observed no significant difference in incubation period across different age groups, including older and younger individuals. These findings suggest a more consistent and brief policy design, for example, using the same period of quarantine regardless of vaccination status or age.

In our data collection and estimation process, we intentionally avoided restrictive case inclusion criteria such as selecting only those with a one-day exposure window, in an attempt to minimize potential selection bias. Given this strategy, coupled with the fact that our data collection period occurred during a stable, low-incidence phase of the outbreak, we believe that our estimate is less prone to the biases associated with selection and dynamic changes.

In our study, we acknowledge several limitations. First, the possibility of other selection biases influencing our results cannot be dismissed. Cases may only recall recent symptoms, potentially overlooking onset dates from a distant past, which could introduce a downward bias. Second, the demographic and social composition of our study cohort may differ from the general population. For instance, an initial outbreak hotspot was among staff at an international airport and a COVID-19 prevention hotel. Third, our meta-analysis did not incorporate a rigorous quality assessment of the included studies. The development of methodologies for quality assessment remains an active field of research. An initial attempt was made by McAlloon et al. [37] for their rapid meta-analysis of pre-Alpha incubation periods, though one of their criteria, favoring a strict selection of cases with a one-day exposure window, may produce downward estimates. Notably, none of the studies included in the meta-analysis of McAlloon et al. met this criterion. Previous studies investigating the 2014-2016 Ebola outbreak in West Africa [38,39] have underscored that including cases with one-day or longer exposure windows may foster more unbiased estimates of the incubation period.

The increased transmissibility of new SARS-CoV-2 variants, along with discontinuation of active case finding and contact tracing efforts, have complicated the estimation of the incubation period and other epidemiological characteristics of SARS-CoV-2. A larger degree of heterogeneity and fewer studies dedicated to estimating the incubation period of Omicron BA.2 and BA.5 variants can be observed (see bottom panel in **Figure 3C**). However, alternative methods that do not rely on contact tracing data have recently been utilized for estimating the incubation period. Ejima et al. [40] previously used the viral load data to estimate the pre-Alpha incubation period. Recently, Russel et al. [29] employed a viral kinetics model to estimating the incubation period of Omicron BA.1 infections with a mean incubation period of 4.9 days (95% CI: 4.2-5.9 days; 48 cases), which is longer than both our estimate and the reported pooled means.

In conclusion, our study provides a detailed estimate of the incubation period for the Omicron BA.1 variant of SARS-CoV-2, by leveraging high-ascertained data from Taiwan during a period of low incidence. Our results hold substantial implications for public health planning and interventions, particularly in the context of evolving SARS-CoV-2 variants. Future studies, including alternative methods that do not rely on contact tracing data, will be essential to continue refining our understanding of the incubation period and other epidemiological parameters for new and emerging SARS-CoV-2 variants.

## Data Availability

All data produced in the present work are contained in the manuscript (Figure 1)

## Acknowledgements

The authors express their gratitude to Natalie M. Linton (California State Department of Health, Richmond, CA, U.S.A.) and Sang Woo Park (Princeton University, Princeton, NJ, U.S.A.) for helpful discussions. They also thank one of the anonymous reviewers of their earlier work [21] for directing them to the quality assessment of meta-analysis studies for infectious diseases conducted by McAloon et al. [37].

## Ethical standards

This study was approved by the Research Ethics Committee of National Taiwan University (202304HM020).

## Data availability statement

All data used for this study can be found at: http://github.com/aakhmetz/OmicronBA1-incper-Taiwan

## Appendix

**Supplementary Figure 1:**
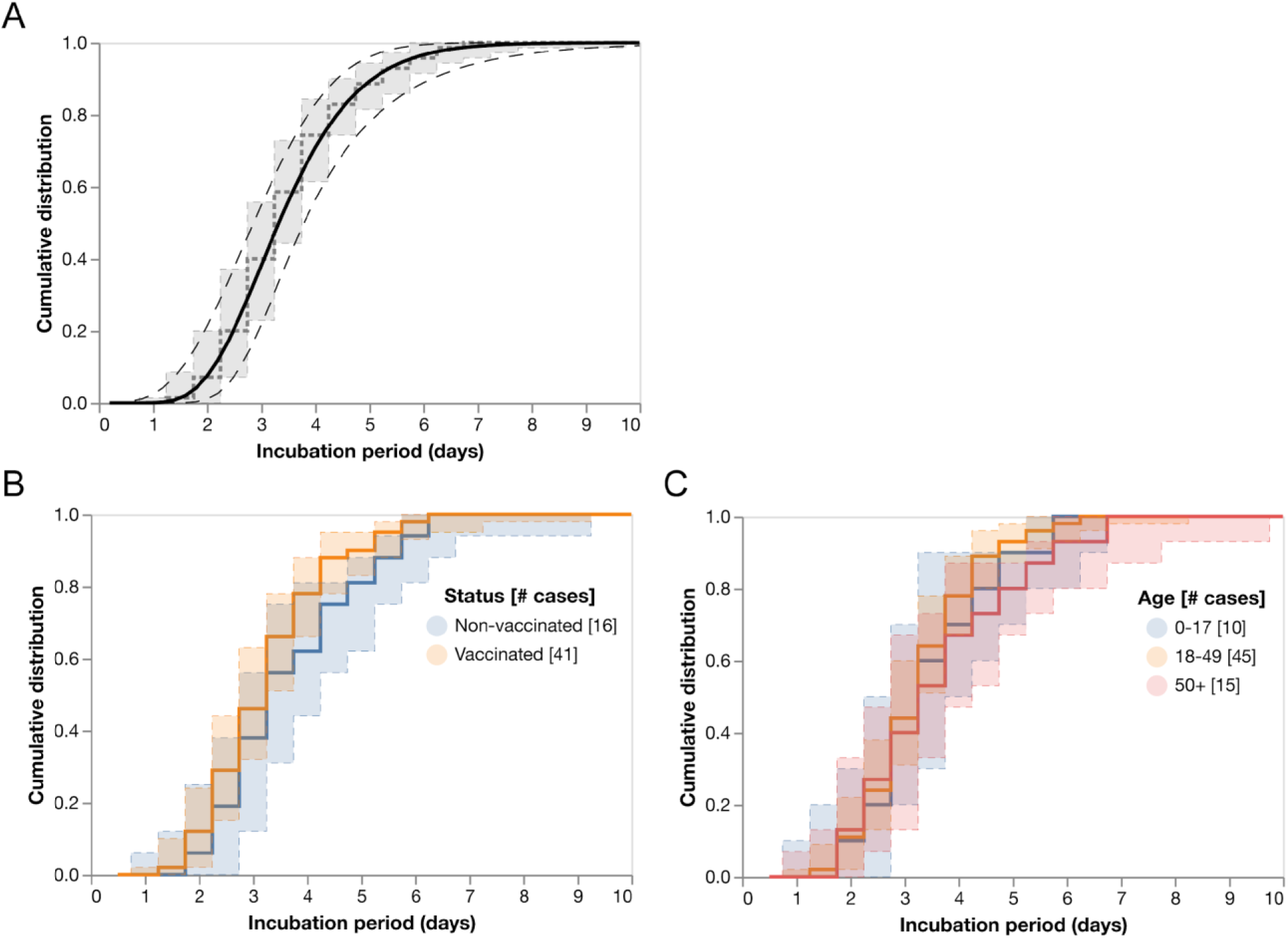
Cumulative distribution function (CDF) of the incubation period (black) compared to the empirical CDF (dashed gray) (**A**); stratified by vaccination status (B); stratified by age-group (C). 95% credible intervals are indicated by shaded areas with their boundaries in dashed lines, except that the shaded area for the present study in (A) is not shown.

